# HISTOPATHOLOGICAL AND IMMUNOHISTOCHEMICAL PROGNOSTIC FACTORS IN HIGH-GRADE NON-ENDOMETRIOID CARCINOMAS OF THE ENDOMETRIUM (HG-NECs). IS IT POSSIBLE TO IDENTIFY SUB-GROUPS AT INCREASED RISK?

**DOI:** 10.1101/2021.05.09.21256904

**Authors:** Valerio Gaetano Vellone, Michele Paudice, Chiara Maria Biatta, Giulia Scaglione, Fabio Barra, Simone Ferrero, Marco Greppi, Laura Paleari, Emanuela Marcenaro, Gyn DMT

**Affiliations:** Dept. of Integrated Diagnostic and Surgical Sciences (DISC), University of Genoa, Italy; Pathology University Unit, San Martino Hospital, Genoa, Italy; Dept. of Neurosciences, Rehabilitation, Ophthalmology, Genetics, Maternal and Child Health (DINOGMI), University of Genoa, Italy; Obstetrics & Gynecology University Unit, San Martino Hospital, Genoa, Italy; Dept. of Experimental Medicine (DIMES) and Centre of Excellence for Biomedical Research (CEBR), University of Genoa; A.Li.Sa., Liguria Region Health Authority, Genoa, Italy

**Keywords:** Endometrial carcinoma, Immunohistochemistry, Prognosis

## Abstract

Endometrial cancer is an emerging disease with an increase in prevalence of aggressive histotypes in recent years. In the present study potential histopathological and immunohistochemical prognostic markers are investigated

Consecutive cases of high grade non-endometrioid carcinoma (HG-NEC) of the endometrium were considered. Each surgical specimen has been routinely processed, the most representative block has been selected for immunohistochemistry and tested for ER, PR, ki67, p53, E-cadherin, β-catenin, Bcl-2 and cyclin D1. For each immunomarker the percentage of positive tumor cells were evaluated (%) and dichotomized as low and high according to the distribution in the study population. Follow-up was collected for disease-free survival (DFS) and overall survival (OS).

33 cases were eligible: 19 resulted FIGO I-II, 14 resulted FIGO III-IV. 12 patients suffered a recurrent disease (mean follow-up 24.6 months); 8 patients died of the disease (mean follow-up 26.6 months).

Women with recurrent disease demonstrated a significant higher bcl2% (35.84±30.96% vs 8.09±11.56% p=0.0032) while DOD patients had higher ki67% (75±13.09% vs 58.6±19.97% p=0.033) and bcl2% of border significance (34.37±34.99% vs 13±17.97% p=0.078).

As expected FIGO III-IV had a worse DFS (HR=3.34; 95%CI:1.1-10.99; p=0.034) and OS (HR=5.19; 95%CI: 1.27-21.14 p= 0.0217). Bcl2-high patients (bcl2>10%) demonstrated a significant worse DFS (HR=9.11; 95%CI: 2.6-32.4; p=0.0006) and OS (HR=7.63; 95%CI:1.7-34; p=0.0084); also PR low patients (PR≤10%) had a significant worse DFS (HR=3.74; 95%CI:1.2-11.9; p=0,02).

HG-NEC represent an heterogeneous group of endometrial aggressive neoplasms with a worrisome prognosis often at an advanced stage at presentation. Bcl2 and PR may represent promising markers to identify a sub-group of patients having an even worse prognosis requiring a careful and close follow-up.

## INTRODUCTION

Endometrial carcinoma (EC) is a very common neoplasm among women, being the sixth cause of cancer in the world, the fourth in the USA and in Italy (5% of all tumors). It counts about 320.000 new cases and 76.000 deaths per year. [1]

Its incidence is higher and still increasing in the western industrialized countries, due to the higher incidence of its risk factors and the longevity of the population. [2]

In Italy it is estimated that 1 every 47 women will develop EC in her life. [3]

75% of EC cases are diagnosed in women older than 50 years old. [4] It also appears that as the age of diagnosis increases, also does tumor aggressiveness, being more frequent TP53 mutations and E-cadherin loss of expression. [5]

EC has been long categorized into two major classes, based on clinical–pathological correlations: type I and type II carcinoma. [6]

EC type I, or endometrioid EC, represents the majority of sporadic endometrial carcinomas (70-80%). It is a moderately indolent tumor that generates after prolonged estrogenic stimulation. EC type II, or non-endometrioid EC, is less frequent (about 10–20% of endometrial carcinomas) but more aggressive and usually not related to estrogen excess or to endometrial hyperplasia. They are typically high-grade carcinomas and include non-endometrioid differentiation, most frequently serous, less frequently clear cell, mixed or undifferentiated. [7]

In this contest high grade non-endometrioid endometrial carcinomas (HG-NECs) constitute the histopathologic manifestation of Type II carcinomas.

Since Bockman’s classification, numerous molecular studies on endometrial cancer have been carried out and dozens of molecular markers have been proposed over the years as prognostic markers. Recently, TCGA (Cancer Genome Atlas Research Network) has performed genomic, transcriptomic and the proteomic characterization of EC using the most modern array and sequencing-based technologies. As result, a new classification dividing ECs in 4 classes has been proposed representing the future in endometrial carcinoma research. [8]

In the current study we have chosen a small panel of molecules valued with immunohistochemistry, previously proposed as prognostic markers in EC, commonly used in daily practice and available in many labs worldwide.

Overall, steroid hormones (mainly estrogen and progesterone) have been considered to play a key role in the pathogenesis of EC, especially in type I carcinoma. Estrogen (ER) and progesterone receptors (PR) are able to influence prognosis and clinical management as well as they correlate with grading and staging. [9]

ERs are expressed in 60-70% of ECs. They have a pivotal role in the carcinogenesis of type I tumors. [10] Conditions resulting in long lasting unopposed exposure to estrogen (obesity, exogenous hormone replacement therapy, polycystic ovary syndrome, anovulation, and type 1/2 diabetes mellitus) can promote the development of atypical endometrial hyperplasia and increase the risk of EC. [11]

The loss of ERα and PR has been correlated with poor survival, whereas expression of ERβ has not shown any clinical pathological correlation. [9] The loss of ERα is associated with high-grade tumors. In contrast, the ERα expression is related to low-grade and low stage of disease.

Progesterone is the physiological estrogen antagonist. [12] It acts by decreasing the risk of developing estrogen-related cancer through several mechanisms, such as reduction of ER and increase in the metabolic inactivation of estrogen. Thus, estrogen-related endometrial hyperplasia can be treated with progestin therapy. [13]

Ki67 is a nuclear antigen expressed by proliferating cells (phases G1, S, G2, mitosis), but absent in resting cells (G0). High Ki67 expression is related to a more aggressive behavior of cancer. [14]

TP53 oncosuppressor gene (chromosome 17) encodes p53 nuclear protein, a transcriptional factor involved in cell cycle arrest and apoptosis. After DNA damage, p53 accumulates and stops the cell cycle through inhibition of cyclin D1 phosphorylation and, if necessary, by promoting apoptosis through interaction with Bax and Apaf 1 proteins. TP53 mutations are typical of EC type II, in particular of serous carcinoma. [15] The majority of TP53 mutations are missense and lead to the loss of oncosuppressor function. In normal cells p53 is rapidly destroyed and cannot be seen by immunohistochemistry (IHC). Missense mutations are clearly visible by IHC because there is nuclear accumulation of aberrant p53: the most common IHC pattern is widespread and intense nuclear positivity. [16]

β catenin is encoded by CTNNB1 gene (chromosome 3) and the protein mediates the link between actin filaments of the cytoskeleton and transmembrane E-cadherin. The IHC nuclear accumulation of β catenin due to gene mutation are significantly more common in EC type I (31-47%) as compared to EC type II (0-3%). On the contrary, E-cadherin mutation is more frequent in EC type 2. Usually, ECs type 1 with CTNNB1 mutation have favorable prognosis and low stage. [17]

E-cadherin is encoded by the CDH1 gene (chromosome 16) and constitutes another adhesion molecule essential for tight junctions between cells. These molecules mediate the connection between cells through a calcium dependent mechanism. [18]

CDH1 is considered an oncosuppressor gene because it controls cell cohesiveness. Low E-cadherin expression is related to major tumor cells exfoliation and high-risk of peritoneal metastasis. E-cadherin mutation is present in 60% of EC type 2 and in 22% of EC type 1, where it is associated with a more aggressive behavior. The partial or total loss of E-cadherin is reported to be associated with adverse prognosis and short survival. [19]

Bcl-2 is a protein with anti-apoptotic activity, which was identified for the first time in non-Hodgkin’s follicular lymphoma. Bcl-2 expression is correlated with many human cancers, including kidney and prostate cancers, thyroid cancer, and non-small cell lung cancer. Loss of BCL2 is associated with independent negative prognostic factors, such as a greater depth of myometrial invasion, aggressive histotype, loss of expression of PR, and advanced FIGO stage at diagnosis. Other studies showed a correlation between loss of BCL2 and risk of lymph node metastasis and recurrence. [20,22]

Cyclin D1,encoded by CCND1, is a proto-oncogene (chromosome 11). Its role is mainly pivotal in the G phase of cell cycle. Cyclin D1 mutation is more typical of EC type 2. [23] Intracytoplasmic protein accumulation, detectable by IHC, has been related to an impairment of proteolytic degradation. [24]

In EC, Cyclin D1 overexpression has a negative prognostic value, and is related with metastatic lymph node involvement. [25]

Rarely β catenin and cyclin D1 are overexpressed together. Some studies showed that cyclin D1 alteration could be an early event in endometrial carcinogenesis, however there is not much difference in its intensity of expression from hyperplasia to EC. [26]

The aim of this study is to identify a sub-group of patients with HG-NECs having a worse prognosis in term of Disease-free Survival (DFS) and Overall Survival (OS) using a limited panel of histopathological and immunohistochemical markers

## MATERIALS AND METHODS

We retrospectively considered all patients treated with radical hysterectomy for endometrial carcinoma in our institution for the period 2013-2018. Only cases with a diagnosis of high grade non-endometrioid carcinoma (HG-NEC) were included. Cases undergone to neoadjuvant chemotherapy, previous hormonal therapy, with incomplete data or follow-up were excluded.

The hysterectomy specimens have been routinely fixed and processed to obtain 3 µm-thick histological sections, finally stained with hematoxylin/eosin. Additional slides have been cut from the most representative paraffin block and tested with a panel of IHC stains including ERα, PR, Ki67, p53, β-catenin, E-cadherin, Bcl-2 and Cyclin-D1. The histopathological examinations have been reported using an institutional protocol.

For IHC we used an automatic immunostainer Benchmark XT (Ventana Medical Systems SA, Strasbourg, France). Antigen-retrieval was obtained with citrate buffer (pH 6) at 90°C for 30 minutes, incubated in primary antibody for 1 hour at 37°C followed by the addition of the polymeric detection system Ventana Medical System Ultraview Universal DAB Detection Kit, counterstained with modified Gill’s hematoxylin and mounted in Eukitt.

The tested antibodies are described in table 1.

**TABLE 1:** LIST OF ANTIBODIES

| Marker | clone | manufacturer | dilution | Low | high |
| --- | --- | --- | --- | --- | --- |
| ER | 6F11 | Ventana | pre-diluted | $\leq 10\%$ | $> 10\%$ |
| PR | 100 | Ventana | pre-diluted | $\leq 10\%$ | $> 10\%$ |
| ki-67 | 30-9 | Ventana | pre-diluted | $< 60\%$ | $\geq 60\%$ |
| p53 | DO-7 | Ventana | pre-diluted | $\leq 10\%$ | $> 10\%$ |
| β-catenin | 14 | Ventana | pre-diluted | $< 70\%$ | $\geq 70\%$ |
| E-cadherin | 36 | Ventana | pre-diluted | $< 70\%$ | $\geq 70\%$ |
| BCL-2 | 124 | Ventana | pre-diluted | $< 60\%$ | $\geq 60\%$ |
| Cyclin D1 | SP4-R | Ventana | pre-diluted | $< 20\%$ | $\geq 20\%$ |

For all the proposed molecular markers, the staining index (SI), accounting the percentage (%) of positive tumor cells, have been evaluated by two pathologists (VGV and MP) working separately and in blind. Any discrepancy has been discussed at a multiheaded microscope to a final decision.

On the base of the distribution in the study population the SIs of each proposed molecular marker have been dichotomized in two discrete categories named low and high according to the cut-off values illustrated in table 1.

The clinical, pathological and IHC data of the patients enrolled in the study were entered in a Microsoft Excel © spreadsheet.

Discrete variables were compared using the χ2 test; continuous variables were compared using Kruskall-Wallis test. Correlations between continuous variables was evaluated with Spearman Rank Correlation. Survival univariate analysis was studied with Kaplan-Meier survival curves. For statistical computation MedCalc© program was used. In all cases a degree of significance of 95% was chosen. In the tables continuous numeric variables are expressed as mean ± standard deviation while continuous variables as the number of observed cases (percentage).

The proposed study has been authorized by the Liguria Regional Ethic Committee (Prot. Pubb.HG-NECs_Vellone2021)

## RESULTS

In the period considered for the purpose of the study a total of 252 EC were collected; 46 had a diagnosis of HG-NEC (18.25%). A total of 33 cases were considered eligible for the aims of the study.

Our study population was composed by elder women (mean 74.12±15.53 years; minimum:53; maximum: 93), often coming to surgery in an advanced stage.

HG-NECs represent a heterogeneous group constituted by different histologic types. The histopathological examination of the surgical specimens frequently showed worrisome features such as infiltrative tumor borders, intratumoral necrosis, and lymph-vascular space invasion. Less frequently moderate/severe desmoplasia or moderate/severe tumor lymphocytic infiltrate were observed. (table 2)

**TABLE 2:**
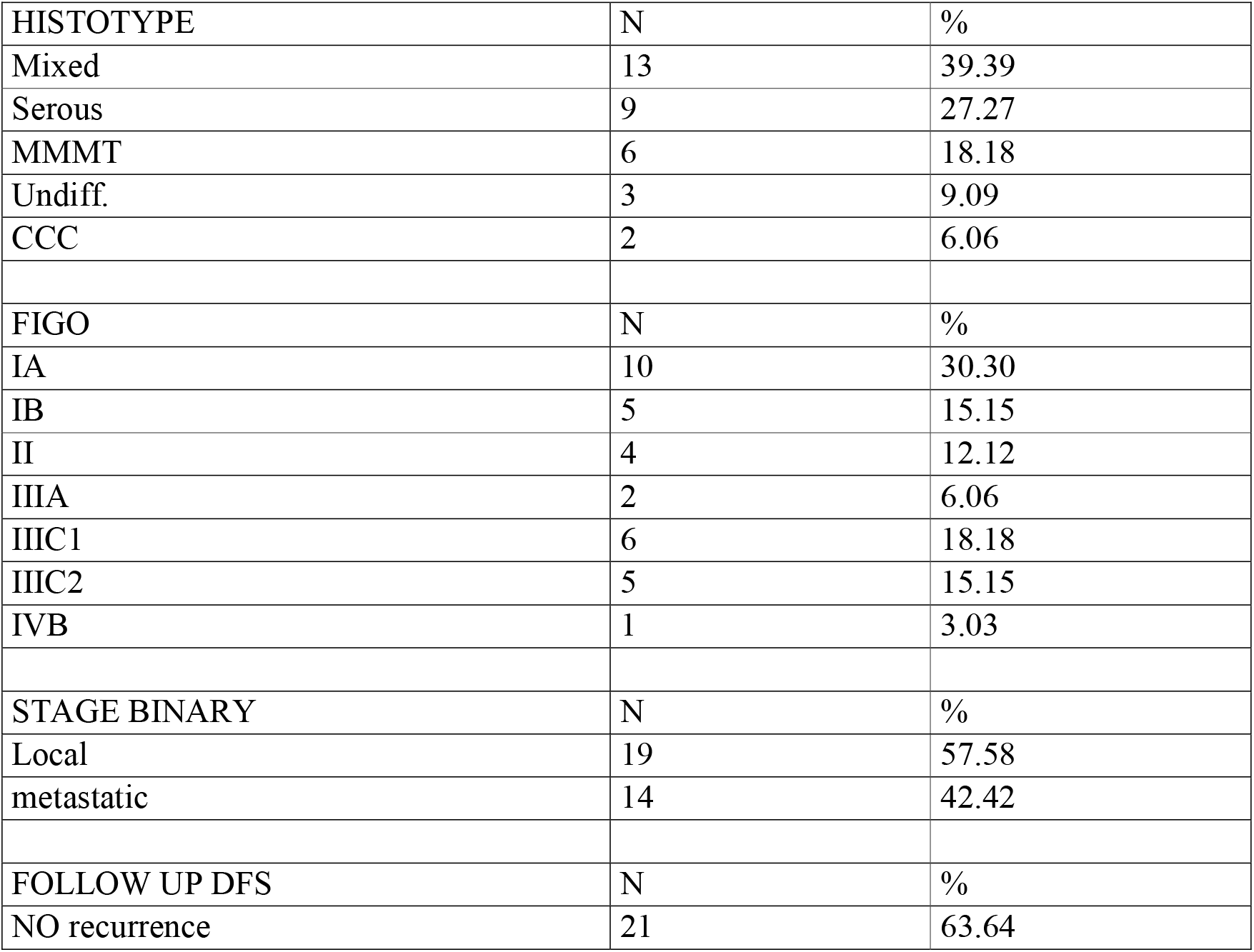
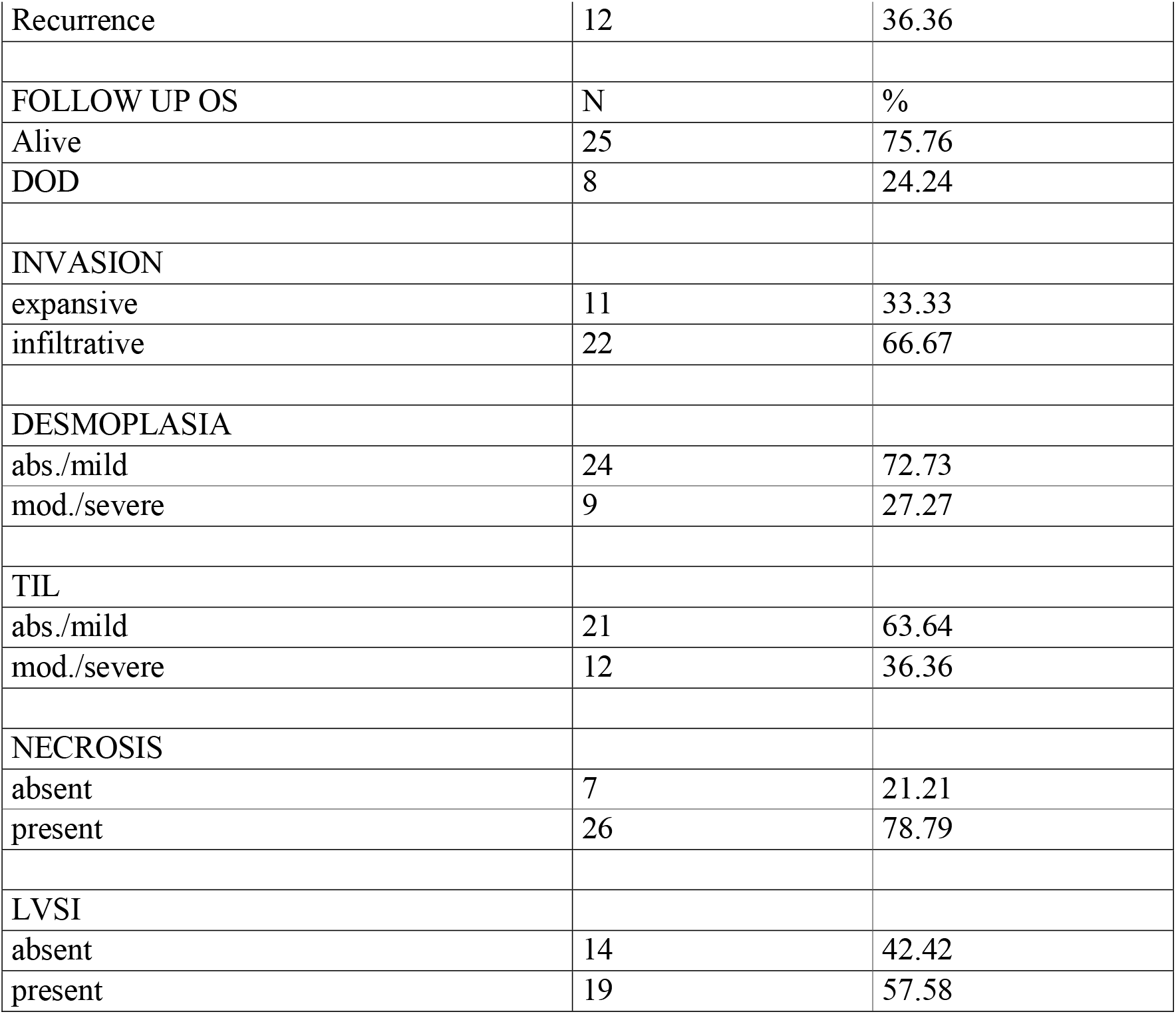
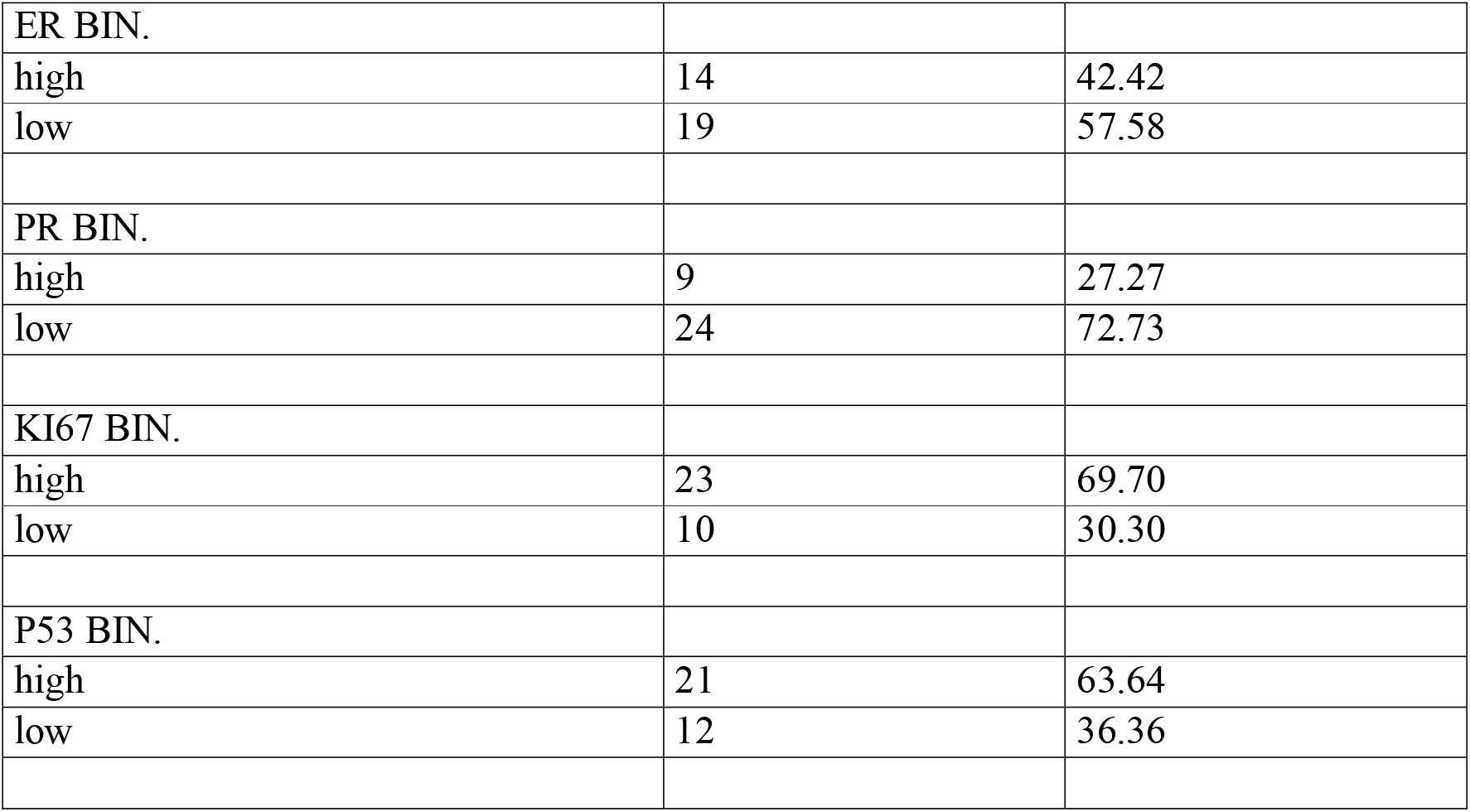
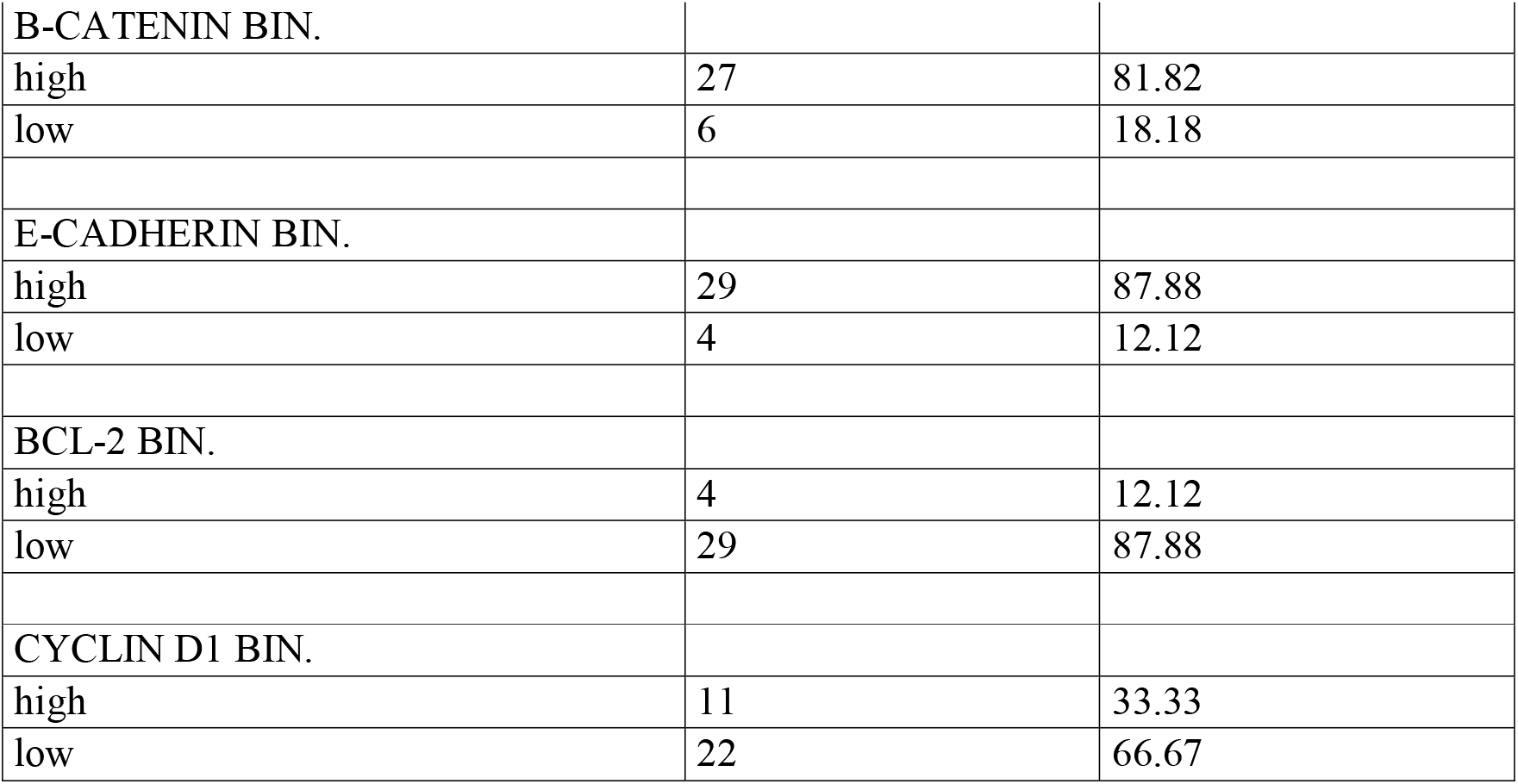
CLINICAL-PATHOLOGICAL AND IMMUNOHISTOCHEMICAL FEAUTURES OF THE STUDY POPULATION. CATEGORICAL VARIABLES. Tab 2.1 Clinical-pathological features Table 2.1 Immunohistochemistry

The immunophenotyping of the neoplasms included in the study showed a fairly wide variability in stain index for all proposed molecular markers: ER and PR were in general low as Bcl-2 and Cyclin D1. while ki67. p53. β-catenin and E-cadherin resulted highly expressed. (table 3)

**TABLE 3.** CLINICAL-PATHOLOGICAL AND IMMUNOHISTOCHEMICAL FEATURES OF THE STUDY POPULATION. CONTINUOS VARIABLES

|  | N | Minimum | Maximum | Mean | Median | SD | RSD | SEM | Normal Distr. |
| --- | --- | --- | --- | --- | --- | --- | --- | --- | --- |
| Age | 33 | 53 | 93 | 74,121 | 76 | 10,532 | 0,1421 | 1,8334 | 0,555 |
| FU DFS Dur. | 33 | 1 | 63 | 24,848 | 22 | 18,3969 | 0,7404 | 3,2025 | 0,1075 |
| FU DOD Dur | 33 | 1 | 63 | 27 | 23 | 18,0624 | 0,669 | 3,1443 | 0,0443 |
| ER% | 33 | 0 | 95 | 25,758 | 5 | 33,7065 | 1,3086 | 5,8675 | 0,0501 |
| PR%_ | 33 | 0 | 60 | 10,788 | 0 | 17,3975 | 1,6127 | 3,0285 | 0,0024 |
| ki67% | 33 | 15 | 90 | 62,576 | 60 | 19,6898 | 0,3147 | 3,4276 | 0,2756 |
| p53% | 33 | 0 | 100 | 51,697 | 60 | 43,8153 | 0,8475 | 7,6273 | <0,0001 |
| B-cat% | 33 | 5 | 100 | 83,939 | 100 | 24,8985 | 0,2966 | 4,3343 | 0,0001 |
| E-cad% | 33 | 20 | 100 | 90 | 100 | 22,3257 | 0,2481 | 3,8864 | <0,0001 |
| Bcl-2% | 33 | 0 | 100 | 18,182 | 10 | 24,4252 | 1,3434 | 4,2519 | 0,0001 |
| Cycl D1% | 33 | 0 | 100 | 21,273 | 15 | 24,1535 | 1,1354 | 4,2046 | 0,0001 |

We observed a fairly strong and significant correlation between ER and PR staining index (rho=0.716; p<0.0001) and between ki67 and p53 (rho=0.541 p=0.0012). (Additional Material 1)

During follow-up (mean DFS follow up duration 24.84±18.39 months; mean OS follow up duration 27±18.06 months). 12 patients (36.36%) recurred after surgery and 8 of them died of the disease (24.24%). as expectable. women with no recurrent disease had a longer follow up (30.28±19.86 months vs 15.33±10.59 months p=0.037).

Considering the proposed markers staining index. women with diseases demonstrated significantly higher levels of Bcl-2 as compared to patients with no recurrent disease (35.84±30.96% vs 8.09±11.56% p=0.0032). No statistically significant differences were demonstrated for the other molecular markers staining indexes. (Additional Material 2)

Patients that died of the disease demonstrated higher levels of ki67 (75±13.09% vs 58.6±19.97% p=0.033) and levels of Bcl-2 tendentially higher (34.37±34.99% vs 13±17.97% p=0.078) than patients alive at end of the follow-up. No statistically significant differences were demonstrated for the other molecular markers staining indexes. (Additional Material 3) On univariate analysis. as expected. patients with metastatic disease at the time of surgery showed a significantly increased risk in both disease recurrence and death by disease. (Table 4.1; Table 5.1; Figure 1)

**TABLE 4:**
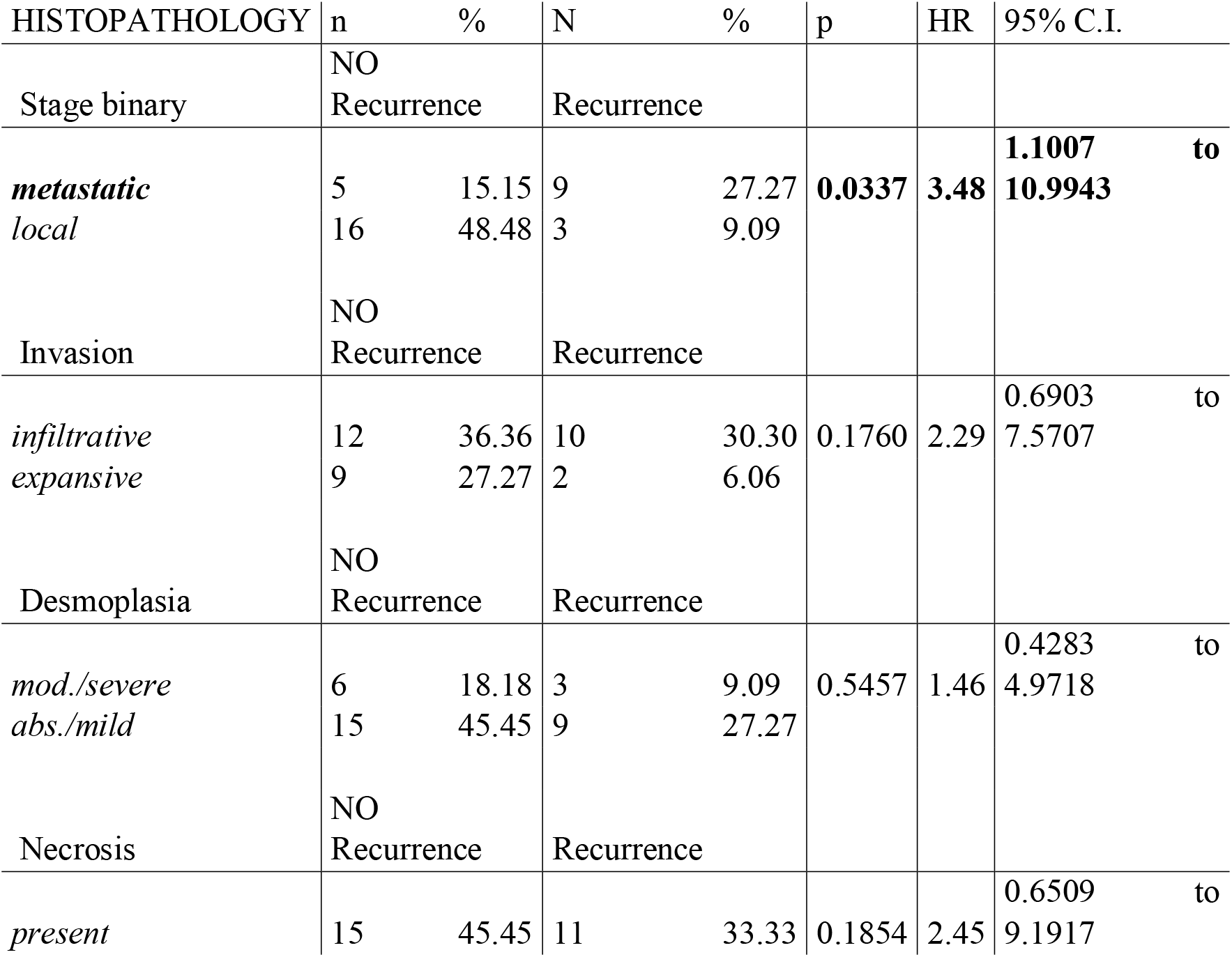
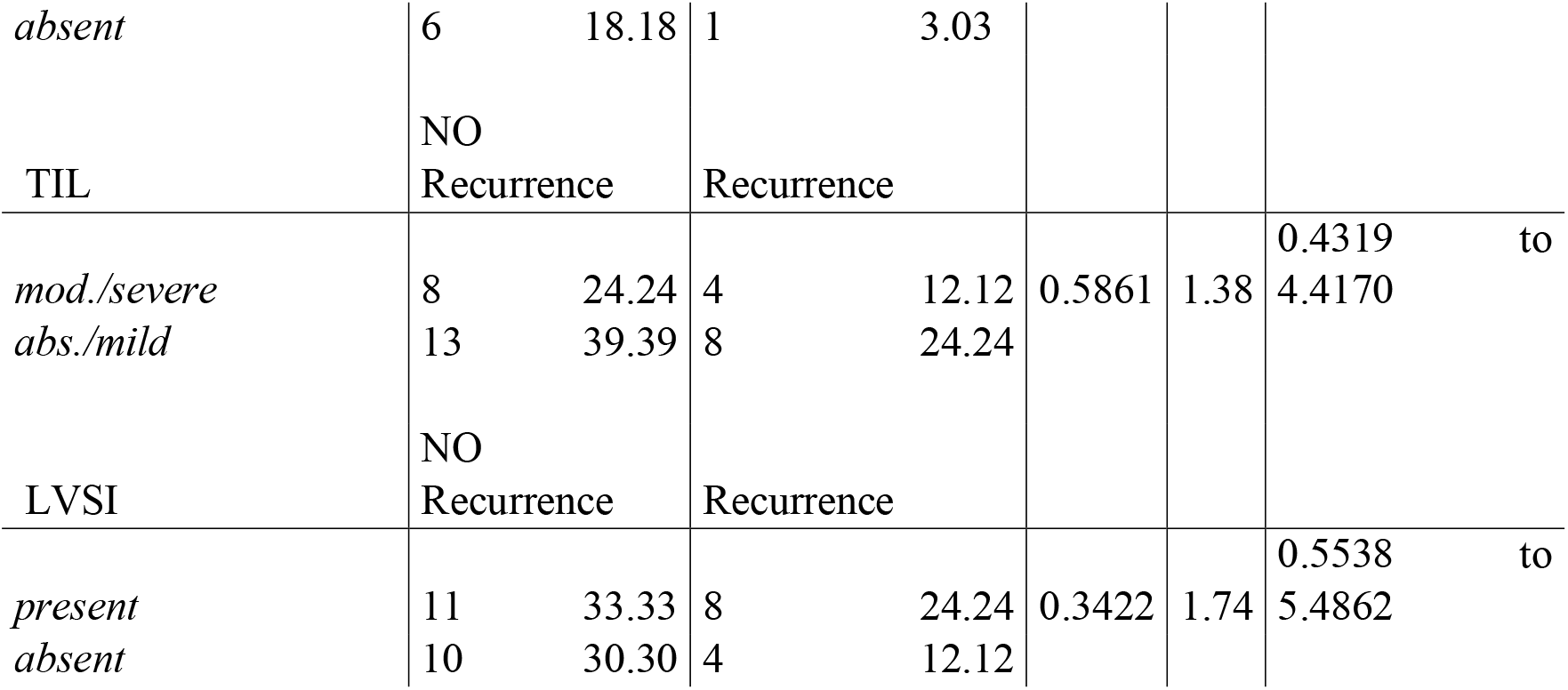
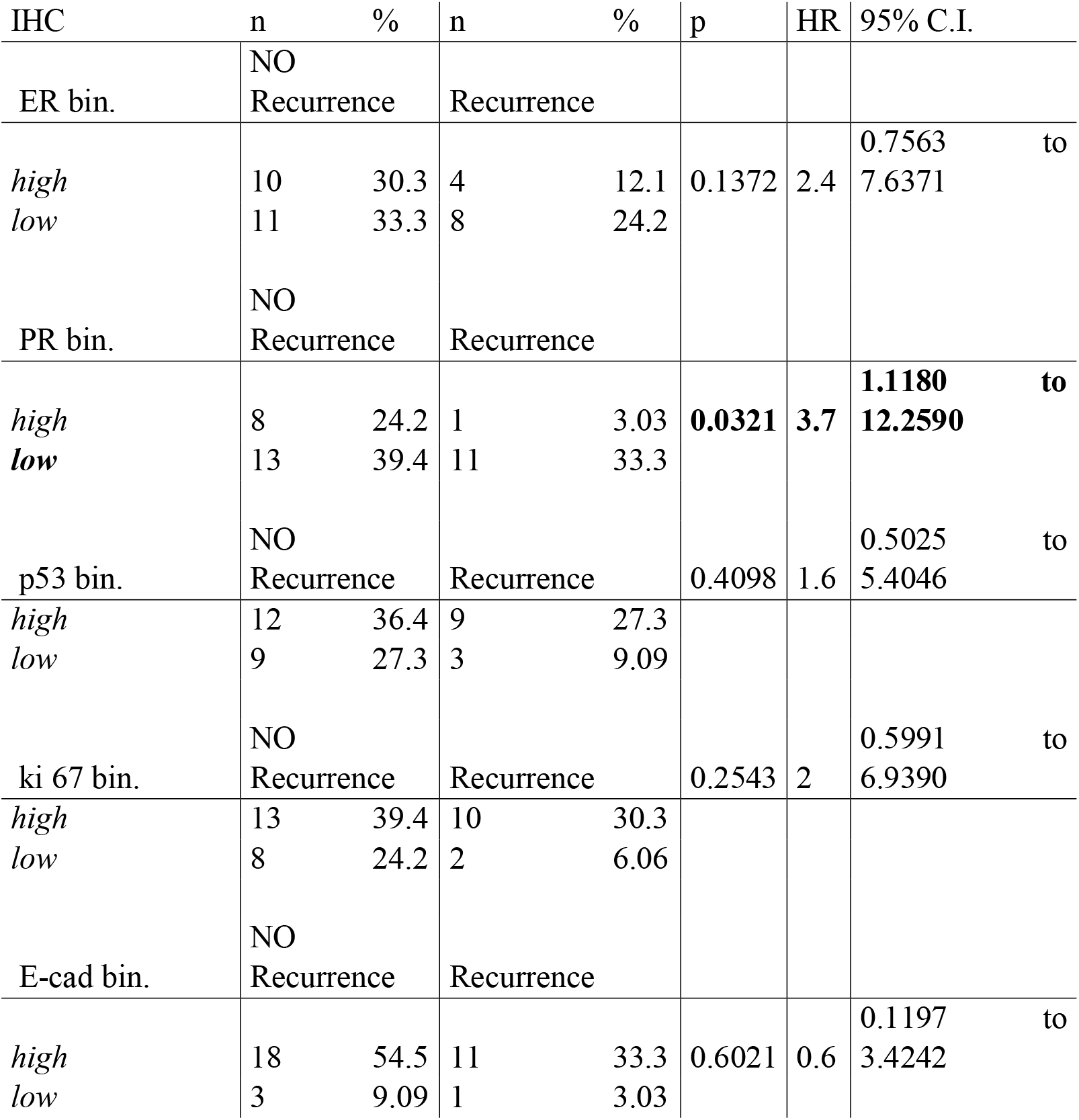
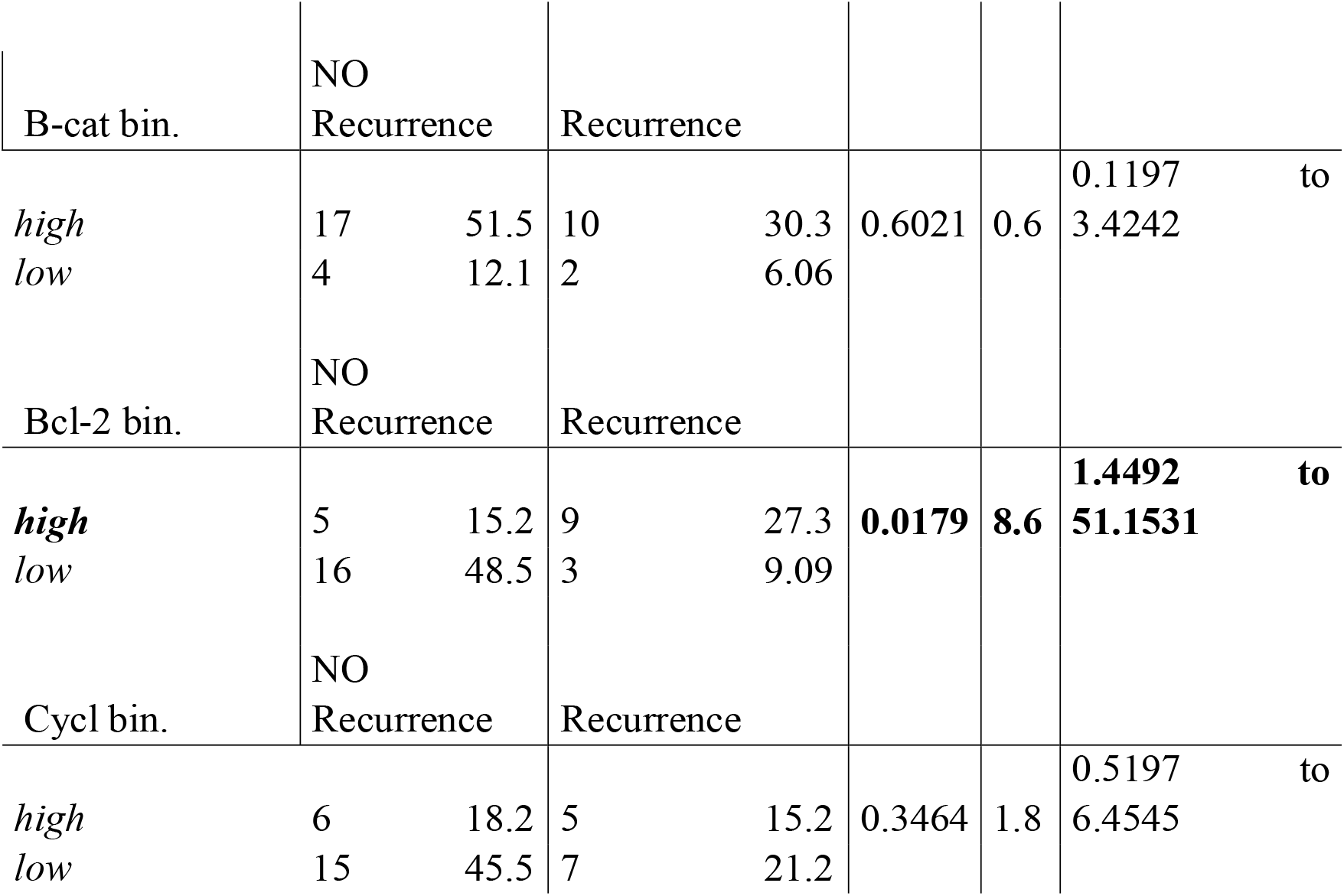
DISEASE FREE SURVIVAL UNIVARIATE ANALYSIS. Table 4.1 HISTOPATHOLOGY Table 4.2 IMMUNOHISTOCHEMISTRY

**TABLE 5:**
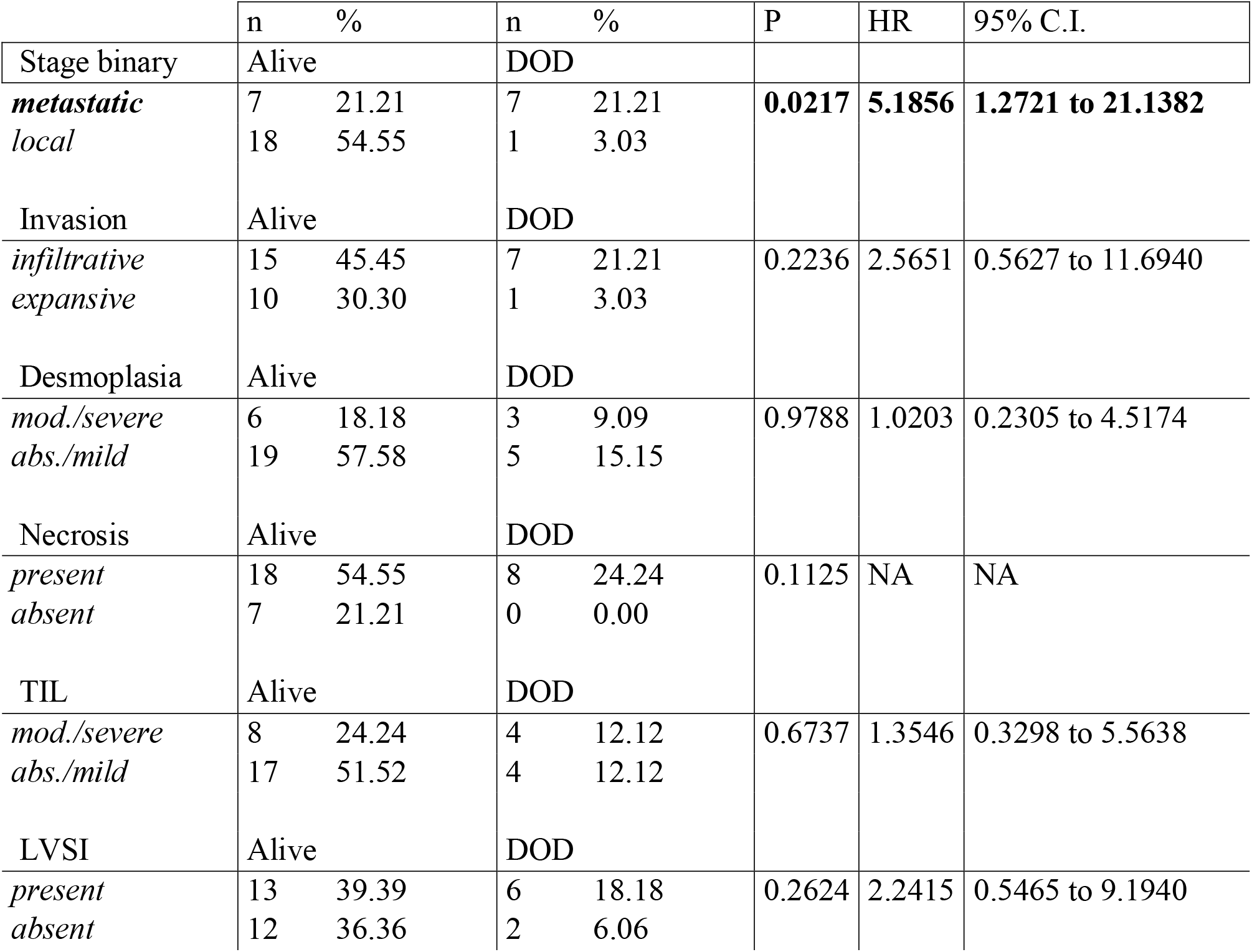
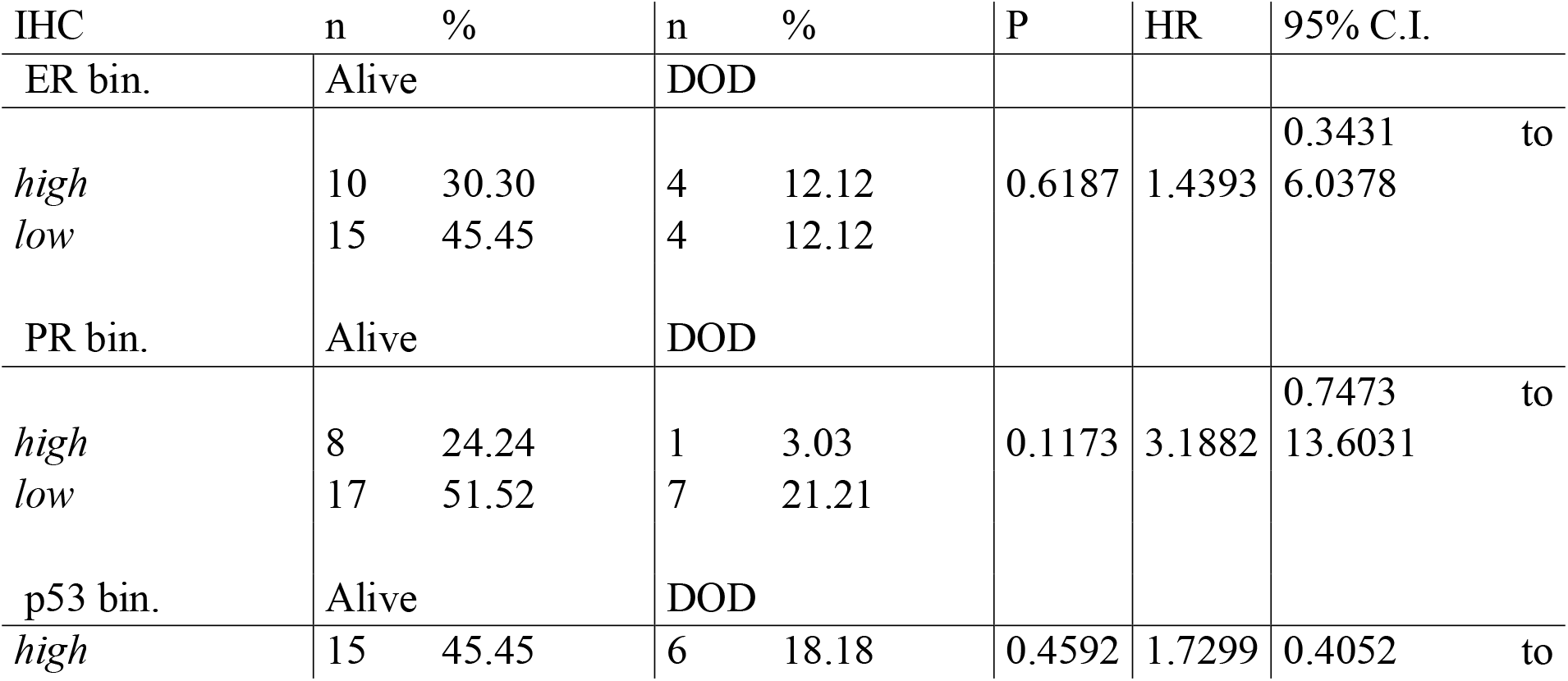
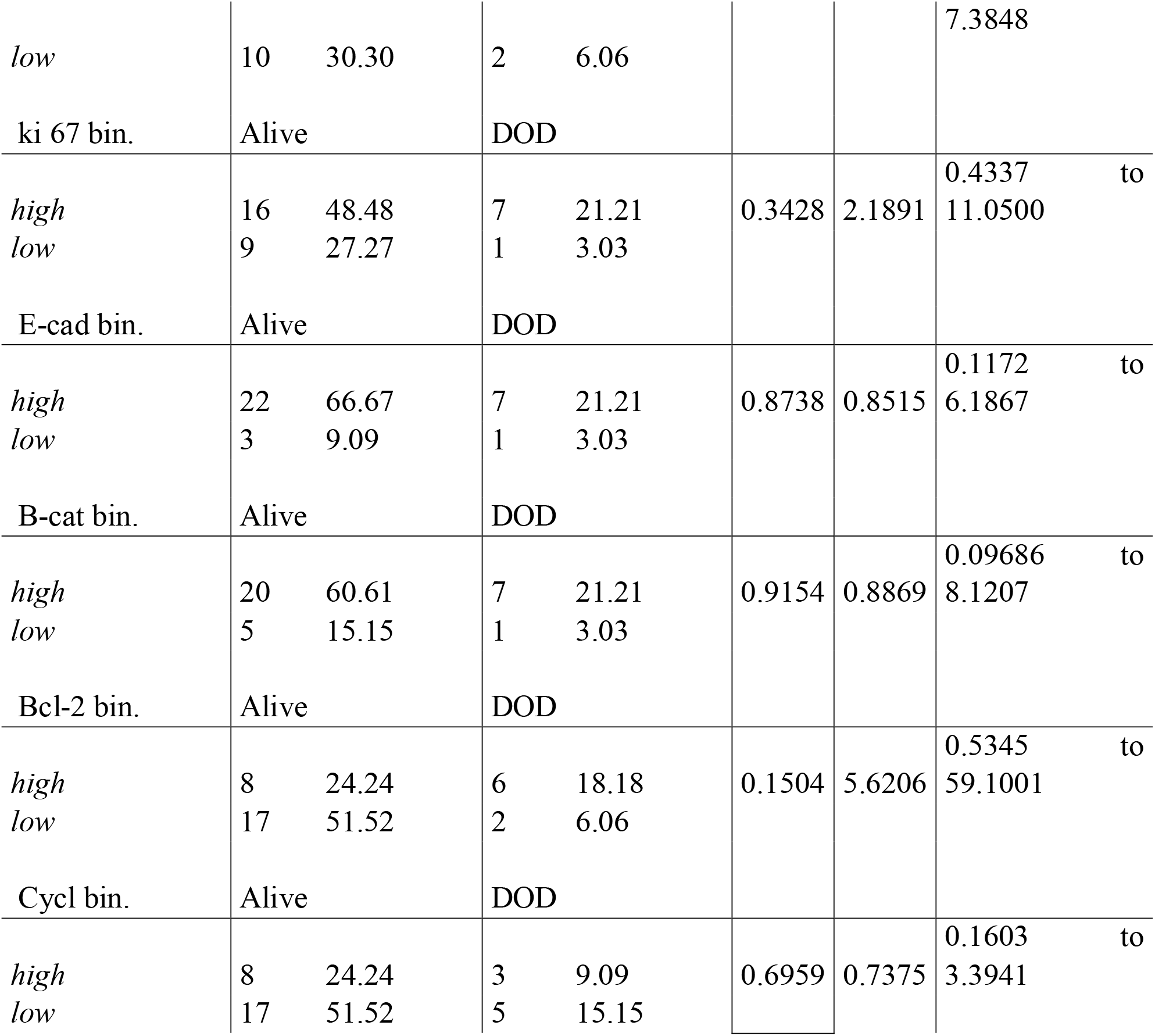
OVERALL SURVIVAL UNIVARIATE ANALYSIS. Table 5.1 HISTOPATHOLOGY Table 5.2 IMMUNOHISTOCHEMISTRY IHC n % n

**FIGURE1:**
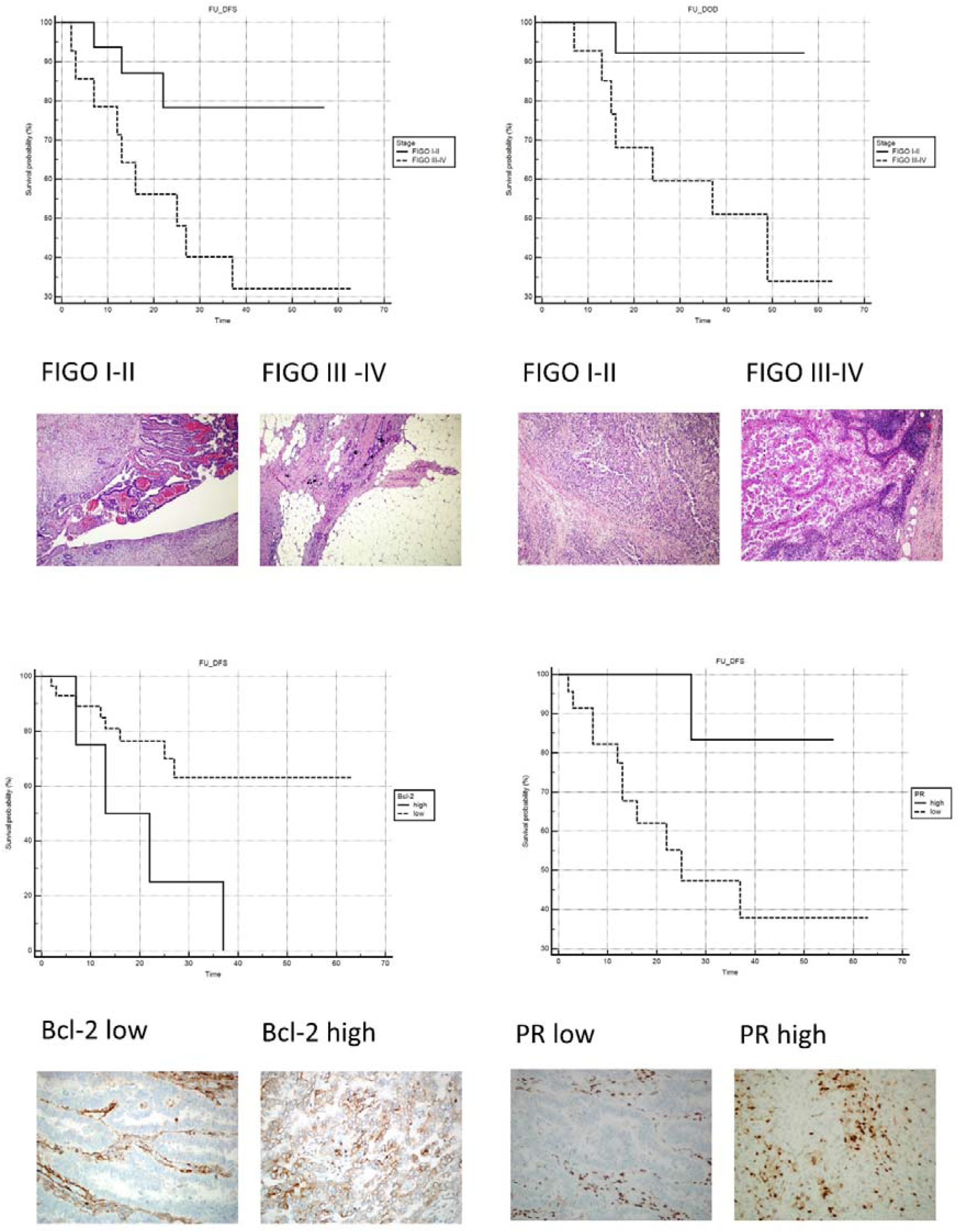

An increased risk of recurrent disease was observed in patients with low PR and high Bcl-2 staining indexes (Table 4.2; Figure1).

The other proposed histopathological and immunohistochemical markers failed to identify a statistically significant risk in term of DFS or OS (Table 4 and 5).

## DISCUSSION

Endometrial carcinoma represents a disease growing in incidence in particular in western countries paralleling the progressive ageing of the population and rising of known risk factors. [27]

In particular the rising of incidence of advanced stages and aggressive histologic types represent a matter of concern for endometrial cancer prevention and treatment.

Only with a better understanding of the molecular events underlying carcinogenesis in the various histotypes of endometrial cancer will it be possible to identify potential prognostic factors and individualized therapy targets.

The recently introduced TGCA classification represent a turn point in endometrial cancer comprehension. Nevertheless, after more than 30 years, the Bokhman and Kurman studies still remain pivotal and relevant. Type I of the traditional classification would encompass cancers belonging to the first three classes of that classification, while Type II of the traditional classification, on the other hand, would encompass tumors of the fourth class. Class 4 Tumors with High Number of Copies are defined as “Serous Like” and are characterized by a high number of aberrations in copy numbers and a low frequency of mutations. They seem to have peculiar mutations frequently involving TP53. FBXW7 and PPP2R1A genes. PTEN and KRAS mutations, typical of low-grade carcinomas with endometrioid histology instead are rare in this kind of tumor. The prognosis of this group appear unfavorable once again. This genomic class includes the majority of serous carcinomas, some of mixed carcinomas and a quarter of endometrioid G3 carcinomas. [28] TGCA classification represents the future but require advanced and expensive molecular techniques currently available in few laboratories and, as a consequence, seldom used as a guide in a real-life clinical setting. Several groups are currently working on surrogate methods to incorporate TGCA findings in clinical practice. [29]

Histopathological examination still represents the first line in daily diagnostic and immunotesting being relatively unexpensive and well established worldwide, they currently represent the cornerstone of any clinical choice. In this contest HG-NECs require a particular attention in search of potential markers of aggressiveness and future targets for individualized therapies. Our series is small but representative of a heterogeneous group of relatively rare malignancies accumulated by aggressive behavior and poor prognosis. [30]

Comparing our results with the current literature was a challenging task, the results often appear contrasting and very few studies are dedicated exclusively to non-endometrioid high grade carcinomas. Many molecular and morphological prognostic factors have been proposed over the years but it is well established that advanced stage and high grade probably are the most important factors affecting the prognosis. [31,32]

An important bias in many studies is represented by grouping all high-grade endometrial carcinomas in single entity. but high-grade endometrial carcinomas (HG-ECs) seems to have profound differences in clinical-pathological and immunophenotype as compared to HG-NECs. [33,34]

HG-NECs constitutes approximately 20% of the EC and affect the elder population. frequently manifesting in an advanced stage of the disease. HG-NECs often present complex histology, more than half of them show more than one histologic type. the largest subgroup was defined as mixed carcinoma, being composed by two epithelial components with at least one of them serous or clear cell. Another sub-group included MMMT which is composed, by definition, by at least two components, one of them epithelial and high grade. In both the sub-group the high-grade epithelial component was prominent accounting at least 30% of the entire tumor mass. We support the hypothesis that this high-grade epithelial component represents the “driving force” of the neoplasm. Pure neoplasms were rarer with incidences in line with the literature. [35]

Although HG-NECs represent a heterogeneous group of tumors they could be grouped by the common finding of worrisome features detectable both in hematoxylin-eosin and with the aid of immunohistochemistry.

Histopathological features. widely considered as markers of aggressiveness such as infiltrative tumor borders. intratumoral necrosis. and lymph-vascular space invasion were a common finding in our series. affecting the majority of the cases. Interestingly other worrisome features such as moderate/severe desmoplasia or moderate/severe tumor lymphocytic infiltrate were observed in a minority of cases. [31]

The wide variability observed on tumor cells morphology has been paralleled in the proposed molecular markers staining indexes.

It is well established how ER is expressed in the majority of endometrial and breast carcinomas and its presence is associated with a less aggressive phenotype [36]. on the contrary and as a general rule. HG-NECs show low levels of steroid hormone receptors. confirming their hormone insensitiveness. PR. in contrast to ER. is suggested to be a more predictive factor of disease-free survival [9] and our findings seems to confirm these observations.

The proliferation index, evaluated with ki67, resulted high in some cases, very high in HG-NECs, confirming their aggressive behavior.

TP53 has a fundamental role in differentiating EC subgroups. The mutation of TP53 represents a crucial event in type II endometrial carcinoma carcinogenesis and progression. It is well reported how its accumulation represents a relevant prognostic factor. [28]

As expected in our study population p53 staining index in general was high, failing to identify sub-groups at increased risk of recurrence or death by disease. It should be noted that immunohistochemistry is able to detect only a part of TP53 mutations.

Even tested adhesion molecules β-catenin and E-cadherin failed to identify sub-groups with increased risk. It is reported how low levels of these molecules are associated with metastatic deposition. [38. 39]. In our study population more than half of the patients with low β-catenin and/or low E-cadherin had FIGO III-IV at surgery.

Bcl-2 is a proto-oncogene that has an anti-apoptotic activity, many regulators of the apoptotic process belong to the same family of bcl-2, which consists of proteins that regulate the permeability of the outer mitochondrial membrane. Some of them have an anti-apoptotic function, such as bcl-2, bcl-xl, and bcl-w, while others have a pro-apoptotic activity, like bax, BAD, bak. and bok. [40,41]

Apoptosis is induced by releasing cytochrome c in the cytosol. with the subsequent activation of caspase 9 and caspase 3. [42]

A theory suggests that Rho proteins may have a role in the activation of bcl-2, bcl-1, and Bid. In fact, the inhibition of Rho decreases the expression of anti-apoptotic proteins and increases the levels of the pro-apoptotic protein Bid. It also induces the release of caspase 9 and caspase 3. [43]

The immunohistochemical staining of bcl-2 in non-neoplastic endometrium has a strong variability; it increases in the proliferative phase and decreases in the secretory phase of the menstrual cycle. In these phases. bcl-2 also plays an important role in regulating the cell differentiation throughout the entire uterine cycle. Some studies have shown that the genes that regulate apoptosis may also be involved in the dysregulation of cell proliferation and death. in the shift from simple to complex hyperplasia. and to adenocarcinoma. [40,41]

The loss of bcl-2 is certainly associated with independent negative prognostic factors, such as deeper myometrial invasion, loss of PR expression, aggressive histotype and advanced FIGO stage. Other studies have shown a correlation between the loss of bcl-2 and the risk of having lymph node metastases and recurrence. [44.45]

In an old study Athanassiadou demonstrated how. on in-print cytological specimens. Bcl-2 expression was associated with a good five year survival. Interestingly also 18 cases of HG-NECs were considered and none of them stained for Bcl-2. [38]

In another, more recent study, Appel et al. failed to find any significant correlation between Bcl-2 expression and histopathologic markers or survival. But again. in this study no distinction between the histologic type is attempted. [22]

Our findings seem to confirm a prognostic role of primary importance for PR and specify the role of Bcl-2 in delimiting a group of patients with greater risk of recurrence. In conclusion we can answer yes to the question in the title. The HG-NECs confirmed their clinical aggressiveness with frequent worrisome aspects. Although marked inter-individual and intratumoral variability has been observed, cases with advanced stage at surgery, low levels of PR and high levels of Bcl-2 have shown a worse DFS. These patients could benefit from a close follow-up with thorough controls and more aggressive treatments.

## Data Availability

Availability of data and material (data transparency): the presented dated are available to asking scholars

## ACKNOWLEDGEMENTS

Special thanks to Giorgia Anselmi. Silvana Bevere. Simona Pigozzi and Laura Zito for technical support

## Notes

Conflicts of interest/Competing interests: no conflicts of interest Availability of data and material (data transparency): the presented dated are available to asking scholars

### Competing Interest Statement

The authors have declared no competing interest.

### Clinical Trial

Retrospective Study

### Funding Statement

Funding: the current work was granted by University of Genoa Research Funds (FRA)

### Author Declarations

Liguria Regional Ethic Committee (Prot. Pubb.HG-NECs_Vellone2021)

