## Additional Material 1 for "HISTOPATHOLOGICAL AND IMMUNOHISTOCHEMICAL PROGNOSTIC FACTORS IN HIGH-GRADE NON-ENDOMETRIOID CARCINOMAS OF THE ENDOMETRIUM (HG-NECs). IS IT POSSIBLE TO IDENTIFY SUB-GROUPS AT INCREASED RISK?"

ADDITIONAL MATERIAL1: CORRELATION TABLE OF CONTINUOS VARIABLES

|  |  | ER%_ | PR%_ | ki67%_ | p53% | β-cat%_ | E-cad%_ | Bcl-2%_ | Cycl-D1% |
| --- | --- | --- | --- | --- | --- | --- | --- | --- | --- |
| ER% | Significance Level P | 0.0066 | 0.1466 | 0.6027 | 0.789 | 0.2118 | 0.8 | 0.0141 | 0.5912 |
|  | Correlation coefficient |  | 0.716 | -0.039 | -0.294 | 0.199 | 0.337 | -0.213 | -0.041 |
|  | Significance Level P |  | <0.0001 | 0.8288 | 0.0966 | 0.2659 | 0.0554 | 0.2334 | 0.8206 |
| PR% | Correlation coefficient | 0.716 |  | -0.054 | -0.236 | 0.14 | 0.343 | -0.254 | -0.136 |
|  | Significance Level P | <0.0001 |  | 0.7656 | 0.186 | 0.4376 | 0.0508 | 0.1544 | 0.4514 |
| ki67% | Correlation coefficient | -0.039 | -0.054 |  | 0.541 | 0.032 | -0.074 | -0.093 | 0.033 |
|  | Significance Level P | 0.8288 | 0.7656 |  | 0.0012 | 0.8589 | 0.6839 | 0.6049 | 0.8553 |
| p53% | Correlation coefficient | -0.294 | -0.236 | 0.541 |  | 0.089 | 0.091 | 0.247 | 0.071 |
|  | Significance Level P | 0.0966 | 0.186 | 0.0012 |  | 0.6217 | 0.6134 | 0.1661 | 0.6927 |
| β-cat% | Correlation coefficient | 0.199 | 0.14 | 0.032 | 0.089 |  | 0.581 | 0.08 | -0.179 |
|  | Significance Level P | 0.2659 | 0.4376 | 0.8589 | 0.6217 |  | 0.0004 | 0.6573 | 0.3199 |
| E-cad% | Correlation coefficient | 0.337 | 0.343 | -0.074 | 0.091 | 0.581 |  | 0.166 | -0.063 |
|  | Significance Level P | 0.0554 | 0.0508 | 0.6839 | 0.6134 | 0.0004 |  | 0.3569 | 0.7271 |
| Bcl-2%_ | Correlation coefficient | -0.213 | -0.254 | -0.093 | 0.247 | 0.08 | 0.166 |  | 0.067 |
|  | Significance Level P | 0.2334 | 0.1544 | 0.6049 | 0.1661 | 0.6573 | 0.3569 |  | 0.7098 |
| Cycl-D1%_ | Correlation coefficient | -0.041 | -0.136 | 0.033 | 0.071 | -0.179 | -0.063 | 0.067 |  |
|  | Significance Level P | 0.8206 | 0.4514 | 0.8553 | 0.6927 | 0.3199 | 0.7271 | 0.7098 |  |
