## Additional Material 2 for "HISTOPATHOLOGICAL AND IMMUNOHISTOCHEMICAL PROGNOSTIC FACTORS IN HIGH-GRADE NON-ENDOMETRIOID CARCINOMAS OF THE ENDOMETRIUM (HG-NECs). IS IT POSSIBLE TO IDENTIFY SUB-GROUPS AT INCREASED RISK?"

ADDITIONAL MATERIAL 2: DFS CONTINUOS VARIABLES

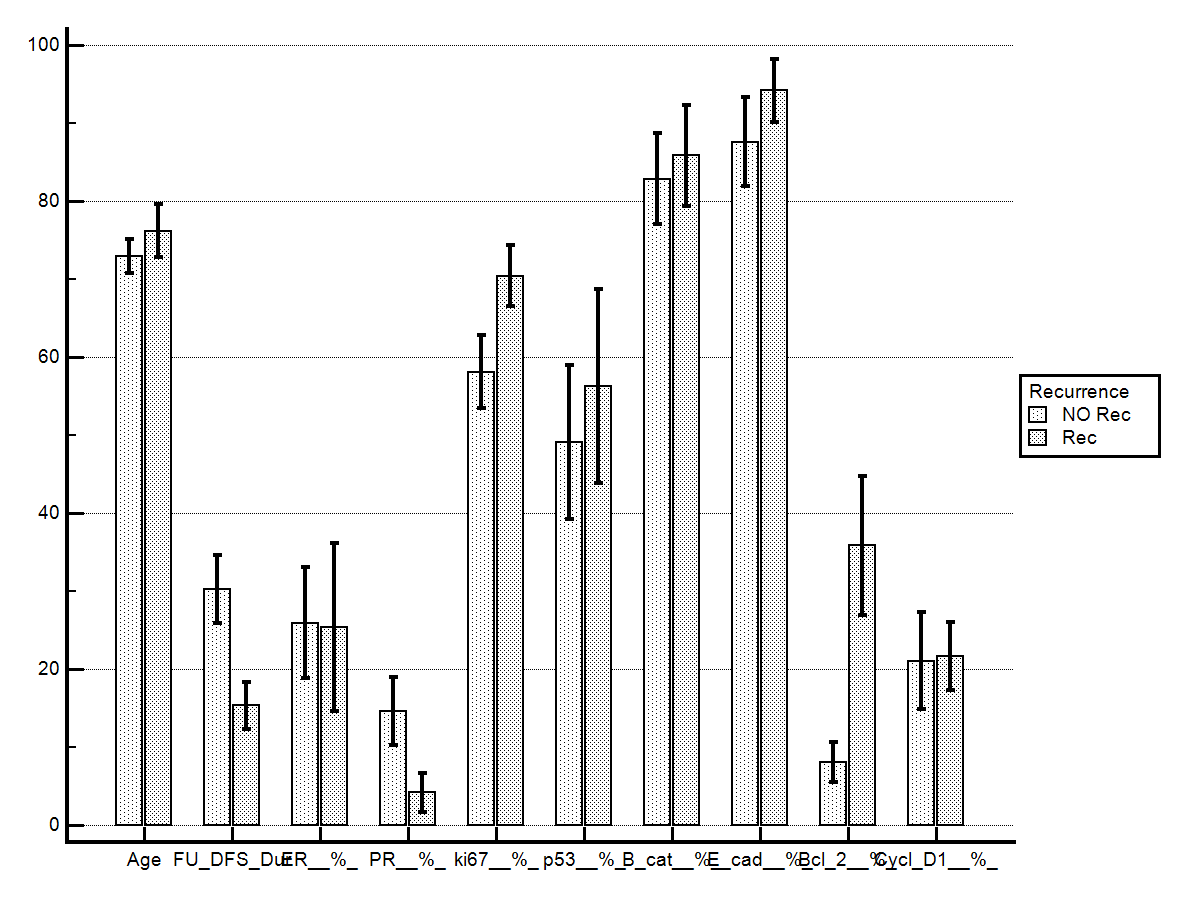

|  | **Age** |  | **FU DFS Dur** |  | **ER %** |  | **PR %** |  | **ki67%** |  |
| --- | --- | --- | --- | --- | --- | --- | --- | --- | --- | --- |
| **FU_DFS** | No Rec | Rec | No Rec | Rec | No Rec | Rec | No Rec | Rec | No Rec | Rec |
| **Mean** | 72,95 | 76,17 | 30,29 | 15,33 | 25,95 | 25,42 | 14,57 | 4,17 | 58,10 | 70,42 |
| **SD** | 9,85 | 11,79 | 19,86 | 10,59 | 32,39 | 37,38 | 20,02 | 8,75 | 21,48 | 13,56 |
| **SEM** | 2,15 | 3,40 | 4,33 | 3,06 | 7,07 | 10,79 | 4,37 | 2,53 | 4,69 | 3,91 |
| **p** | 0,30 |  | **0,03767** |  | 0,84 |  | 0,24 |  | 0,13 |  |
|  | **p53%** |  | **B-cat%** |  | **E-cad%_** |  | **Bcl-2%** |  | **Cycl D1%** |  |
| **FU_DFS** | No Rec | Rec | No Rec | Rec | No Rec | Rec | No Rec | Rec | No Rec | Rec |
| **Mean** | 49,10 | 56,25 | 82,86 | 85,83 | 87,62 | 94,17 | 8,10 | 35,83 | 21,05 | 21,67 |
| **SD** | 45,05 | 43,12 | 26,72 | 22,34 | 26,01 | 13,79 | 11,56 | 30,96 | 28,42 | 15,13 |
| **SEM** | 9,83 | 12,45 | 5,83 | 6,45 | 5,68 | 3,98 | 2,52 | 8,94 | 6,20 | 4,37 |
| **p** | 0,64 |  | 0,72 |  | 0,34 |  | **0,00327** |  | 0,32 |  |
