## Additional Material 3 for "HISTOPATHOLOGICAL AND IMMUNOHISTOCHEMICAL PROGNOSTIC FACTORS IN HIGH-GRADE NON-ENDOMETRIOID CARCINOMAS OF THE ENDOMETRIUM (HG-NECs). IS IT POSSIBLE TO IDENTIFY SUB-GROUPS AT INCREASED RISK?"

ADDITIONAL MATERIAL 3: OS CONTINUOS VARIABLES

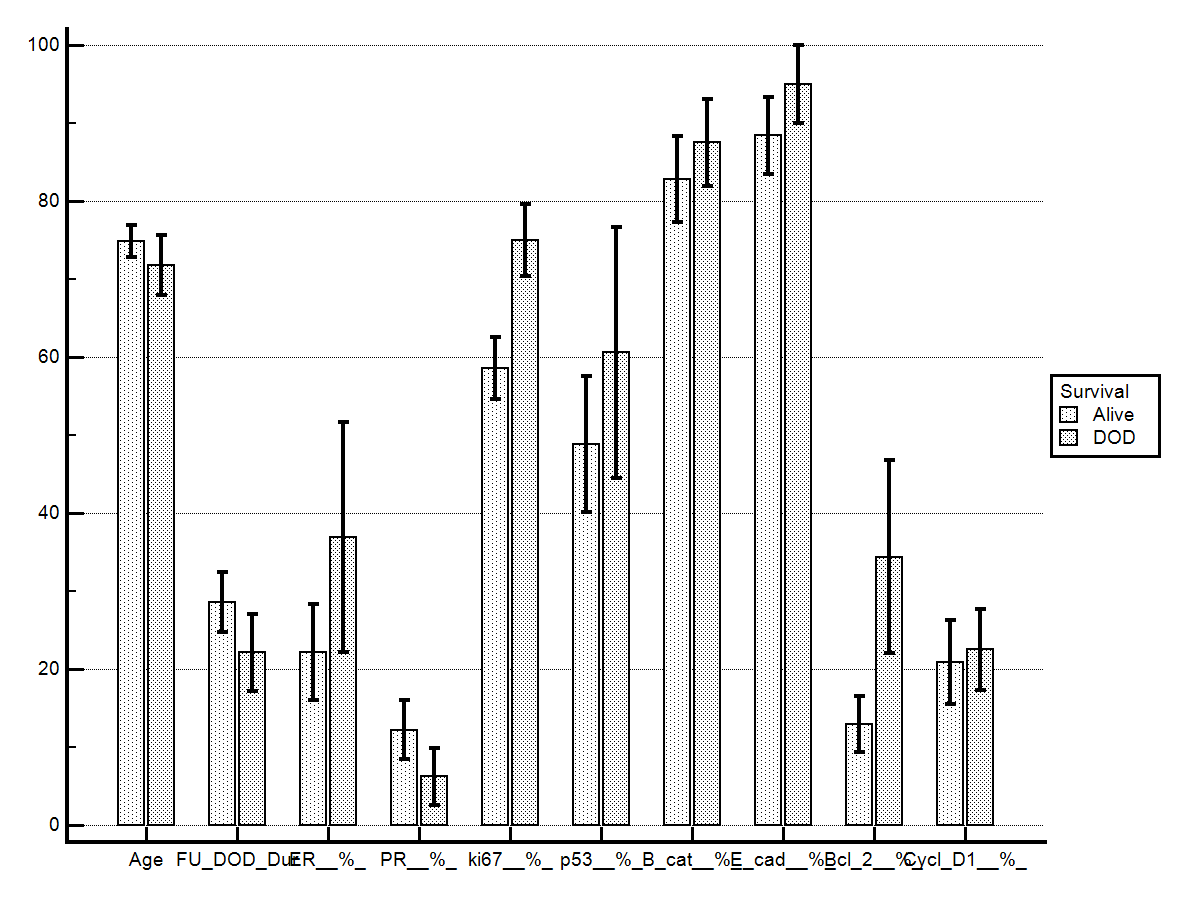

|  | **Age** |  | **FU DOD_Dur** |  | **ER %** |  | **PR %** |  | **ki67%** |  |
| --- | --- | --- | --- | --- | --- | --- | --- | --- | --- | --- |
| **FU_DOD** | Alive | DOD | Alive | DOD | Alive | DOD | Alive | DOD | Alive | DOD |
| **Mean** | 74,88 | 71,75 | 28,56 | 22,125 | 22,2 | 36,875 | 12,24 | 6,25 | 58,6 | 75 |
| **SD** | 10,52 | 10,925 | 19,1531 | 14,0655 | 30,8923 | 41,6565 | 19,0728 | 10,3 | 19,975 | 13,1 |
| **SEM** | 2,103 | 3,8626 | 3,8306 | 4,9729 | 6,1785 | 14,7278 | 3,8146 | 3,63 | 3,995 | 4,63 |
| **p** | 0,69 |  | 0,50114 |  | 0,36531 |  | 0,981363 |  | **0,033036** |  |
|  | **p53%** |  | **B-cat%** |  | **E-cad%_** |  | **Bcl-2%** |  | **Cycl D1%** |  |
| **FU_DOD** | Alive | DOD | Alive | DOD | Alive | DOD | Alive | DOD | Alive | DOD |
| **Mean** | 48,84 | 60,625 | 82,8 | 87,5 | 88,4 | 95 | 13 | 34,4 | 20,88 | 22,5 |
| **SD** | 43,81 | 45,547 | 27,3511 | 15,8114 | 24,3977 | 14,1421 | 17,9699 | 35 | 26,7337 | 14,6 |
| **SEM** | 8,763 | 16,103 | 5,4702 | 5,5902 | 4,8795 | 5 | 3,594 | 12,4 | 5,3467 | 5,18 |
| **p** | 0,551 |  | 0,981935 |  | 0,32181 |  | 0,078521 |  | 0,269308 |  |
